# Auricular Vagus Nerve Stimulation Mitigates Inflammation and Vasospasm in Subarachnoid Hemorrhage: A Randomized Trial

**DOI:** 10.1101/2024.04.29.24306598

**Authors:** Anna L Huguenard, Gansheng Tan, Dennis J Rivet, Feng Gao, Gabrielle W Johnson, Markus Adamek, Andrew T Coxon, Terrance T Kummer, Joshua W Osbun, Ananth K Vellimana, David D Limbrick, Gregory J Zipfel, Peter Brunner, Eric C Leuthardt

**Author notes:** Corresponding Author: Corresponding Author: Eric Leuthardt, Washington University in St. Louis 660 S. Euclid Campus Box 8057 St. Louis, MO 63110 Preliminary findings presented at the American Association of Neurological Surgeons meeting (Philadelphia, PA 04/2022), Academy of Neurological Surgeons meetings (Colorado Springs, CO 10/2022), and Society of NeuroInterventional Surgery Annual Meeting (San Diego, CA 08/2023), and the Southern Neurosurgical Society Meeting (Orlando, FL 03/2024) Twitter handles: @WashUNeurosurg @ahuguena @leuthardt. <u>Non-Standard Abbreviations and Acronyms:</u> CSD: cortical spreading depolarizations, CSF: cerebrospinal fluid, EVD: external ventricular drain, SAH: subarachnoid hemorrhage, taVNS: transauricular vagus nerve stimulation, TENS: transcutaneous electrical nerve stimulation, VNS: vagus nerve stimulation, taVNS: transauricular vagus nerve stimulation.

## Abstract

**Background:** Inflammation contributes to morbidity following subarachnoid hemorrhage (SAH). Transauricular vagus nerve stimulation (taVNS) offers a noninvasive approach to target the inflammatory response following SAH.

**Methods:** In this prospective, triple-blinded, randomized, controlled trial, twenty-seven patients were randomized to taVNS or sham stimulation. Blood and cerebrospinal fluid (CSF) were collected to quantify inflammatory markers. Cerebral vasospasm severity and functional outcomes (modified Rankin Scale, mRS) were analyzed.

**Results:** No adverse events occurred. Radiographic vasospasm was significantly reduced (p = 0.018), with serial vessel caliber measurements demonstrating a more rapid return to normal than sham (p < 0.001). In the taVNS group, TNF-α was significantly reduced in both plasma (days 7 and 10) and CSF (day 13); IL-6 was also significantly reduced in plasma (day 4) and CSF (day 13) (p < 0.05). Patients receiving taVNS had higher rates of favorable outcomes at discharge (38.4% vs 21.4%) and first follow-up (76.9% vs 57.1%), with significant improvement from admission to first follow-up (p = 0.014), unlike the sham group (p = 0.18). The taVNS group had a significantly lower rate of discharge to skilled nursing facility or hospice (p = 0.04).

**Conclusion:** taVNS is a non-invasive method of neuro- and systemic immunomodulation. This trial supports that taVNS following SAH can mitigate the inflammatory response, reduce radiographic vasospasm, and potentially improve functional and neurological outcomes.

Clinical Trial Registration: https://clinicaltrials.gov/ct2/show/NCT04557618

## Introduction

Subarachnoid hemorrhage (SAH) resulting from a ruptured aneurysm accounts for 7% of all strokes worldwide, with 40% of patients suffering permanent disability^1^. Secondary injury is a major driver of morbidity following SAH, as mediated by early brain injury, cerebral vasospasm, delayed cortical ischemia, and chronic hydrocephalus^2^.

Inflammation is a key factor driving the morbidity associated with SAH. Following SAH, blood within the subarachnoid space triggers local and systemic inflammatory responses with increases in the inflammatory markers IL-6^3–5^ and TNF-α^6^ both systemically and centrally. Notably, these increased inflammatory markers are correlated with adverse sequelae from SAH, including the risk for vasospasm, cerebral edema, hydrocephalus, and poor overall patient outcome^3,6^.

Despite understanding the role of inflammation following SAH, an effective method to modulate the deleterious inflammatory response in patients following SAH is lacking. Pharmacologic approaches that target anti-inflammatory pathways have been unsuccessful in clinical trials^7,8^.

Vagus nerve stimulation (VNS) provides a novel, non-pharmacologic approach to systemic immunomodulation. Studies have demonstrated that VNS reduces systemic inflammatory markers^9^ and has had early success treating inflammatory conditions such as arthritis^10^, sepsis^11^, and inflammatory bowel disease^12^. Early evidence has also emerged that non-invasive approaches via transcutaneous auricular vagus nerve stimulation (taVNS) can accomplish similar effects^13^. Thus, our hypothesis was that implementing taVNS in the acute period following spontaneous SAH would attenuate the expected inflammatory response to hemorrhage and curtail clinical morbidity.

Here, we report results from the *Non-invasive Auricular Vagus nerve stimulation for Subarachnoid Hemorrhage* (NAVSaH) randomized control trial^14^. The primary aims of this trial were to determine if taVNS following SAH reduces TNF-α in the plasma and CSF and reduces the rate of radiographic vasospasm. Exploratory analyses also evaluated the change in clinical outcomes of patients via blinded assessment with modified Rankin Scale scores (mRS) at discharge and follow-up. In aggregate, the findings support taVNS as a potential novel modality to treat SAH-induced inflammation and associated clinical sequelae.

## Methods

A full outline of the Non-invasive Auricular Vagus nerve stimulation for Subarachnoid Hemorrhage (NAVSaH) trial has been previously published^14^.

### Trial Design

This study was a prospective, triple-blind, randomized control trial with two study arms, with assessment based on intention to treat with regard to the assigned treatment arm. Reporting followed the Consolidated Standards of Reporting Trials (CONSORT) guidelines^15^. This study was approved by the Washington University School of Medicine institutional review board. All participants enrolled in the study had written informed consent. All procedures followed were in accordance with institutional guidelines. (**Fig. 1)**

**Figure 1.**
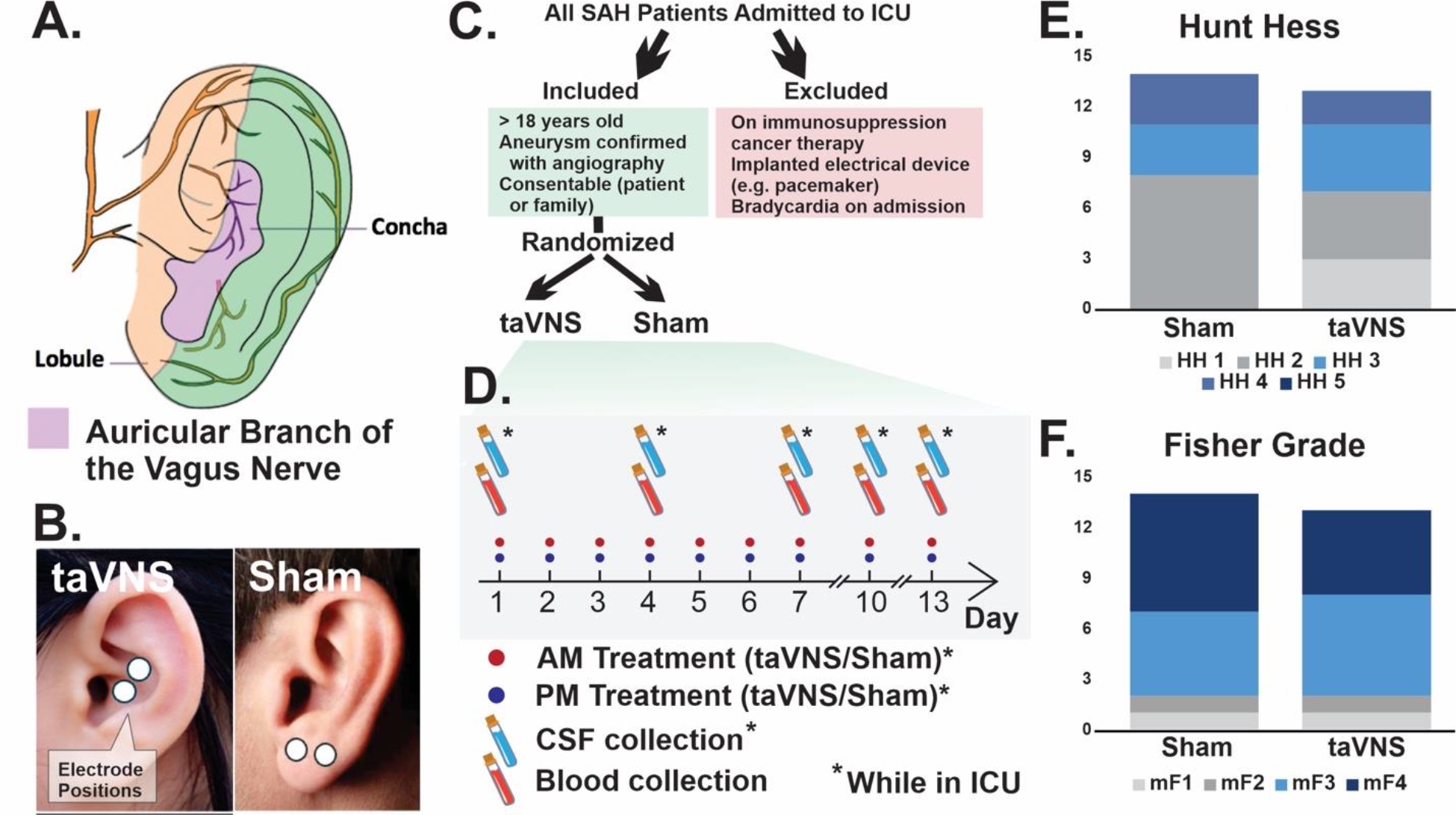
Methods and trial design. Anatomy and cutaneous innervation of the outer ear by the auricular branch (**A**). Position of the electrodes for patients assigned to the vagus nerve stimulation (left) and sham (right) treatment groups (**B**). Patients were screened for adherence to inclusion/exclusion criteria (**C**), and following randomization underwent treatment sessions and blood/cerebrospinal fluid collection every 3 days (**D**). Patients in both treatment groups had similar distributions of Hunt and Hess Scores (**E**) and modified Fisher Grade scores (**F**) on presentation. taVNS= transauricular vagus nerve stimulation, SAH= subarachnoid hemorrhage, HH= Hunt and Hess, mF= modified Fisher

### Participants

Adult patients admitted to Barnes Jewish Hospital following an acute, spontaneous (non-traumatic) SAH were screened for enrollment into the trial. Inclusion criteria included 1) patients with CT-identified, non-perimesencephalic SAH, 2) age ≥ 18 years, and 3) the patient or legal representative able to give consent for enrollment. Patients were excluded if they were 1) less than 18 years old, 2) taking an immunomodulatory or immunosuppressive medication, 3) receiving ongoing cancer therapy, 4) had an implanted electrical device such as a pacemaker, 5) bradycardic on admission (heart rate < 40 beats per minute for greater than 10 minutes), 6) positive for the COVID-19 virus, or 7) at risk of imminent death or were not being offered traditional interventions because of poor prognosis. All candidates were screened based on inclusion and exclusion criteria, consent was obtained, and subjects were enrolled within 24 hours of presentation to our hospital. (**Fig.1 C**)

### Randomization

All identified subjects or their legal medical representative underwent informed consent, and randomization occurred after consent and trial enrollment was complete. Subjects were allocated to treatment arm using a computer-generated randomization sequence with next assignment concealed prior to time of randomization. Consequent eligible patients were allocated strictly in sequence. Patients were randomized to receive either taVNS or sham treatment at a 1:1 ratio.

### Blinding

The patient/family, the care providers and medical team, and the outcomes assessors were blinded to the treatment arm in which the patients were enrolled. Both treatment arms included the application of ear clip electrodes for the same duration and at the same time intervals, and stimulation parameters used in the taVNS arm were sub-sensory, ensuring the patient and casual observer remained blinded to the treatment arm.

### Interventions

Following enrollment and randomization into a treatment arm, initial samples of blood and CSF (if the patient had an external ventricular drain in place) were collected. The patient began either taVNS or sham stimulation twice daily during their stay in the intensive care unit. All patients were fitted with a portable TENS (transcutaneous electrical nerve stimulation) unit connected to two ear clips applied to the left ear during treatment periods. For VNS treatment, these ear clips were placed along the concha of the ear; for sham treatments, the clips were placed along the ear lobe to avoid stimulation of the auricular vagus nerve from tactile pressure alone in the absence of current^16^ (**Fig. 1 A&B**). Stimulation parameters were selected based on prior studies that sought to maximize vagus nerve stimulation while avoiding the perception of pain^16,17^. Stimulation parameters used in this trial for the taVNS arm were 20 minutes in duration, frequency of 20 Hz, 250µs pulse width, and an intensity of 0.4mA. Sham treatments involved no electrical current, and also had a 20-minute duration.

### Outcomes

All outcomes were assessed based on the intention to treat with regard to the assigned treatment arm.

### Impact of taVNS Following SAH on the Inflammatory Markers TNF-α and IL-6

The primary outcome assessed was the change in the key inflammatory marker TNF-α in the plasma and CSF in SAH patients undergoing the taVNS versus sham treatment. IL-6 was also evaluated as a secondary outcome in an exploratory manner. Blood samples and CSF samples, when a ventriculostomy was in place, were collected every three days throughout patient hospitalization following acute SAH. Baseline blood and CSF samples were collected prior to the first treatment session. Blood and CSF samples were collected and processed immediately. See **Supplemental Materials** for details of specimen processing.

### Impact of taVNS on SAH-induced Angiographic Vasospasm

The primary outcome assessed was the difference in the presence of moderate or severe radiographic vasospasm between the taVNS and sham stimulation groups, as assessed by blinded clinician assessment and quantitatively via measurement of vessel caliber on serial exams. In addition to initial diagnostic testing, all patients underwent a repeat CT or catheter angiogram approximately seven days after admission per the institution’s protocol. Further vascular imaging was also performed if there was a clinical concern for clinical vasospasm or stroke. For both planned and indicated imaging sessions, each vascular imaging study was reviewed by a neuroradiologist or endovascular neurointerventionalist blinded to treatment arm, who described the overall imaging study as it related to vasospasm as none, mild, moderate, or severe to account for proximal vessel caliber and overall/distal perfusion.

To assess radiographic vasospasm more quantitatively, all radiological images were reviewed by a single physician reviewer (DJR) blinded to patient identity, cohort assignment, and indication for the imaging study. The reviewer was fellowship-trained in neurointerventional radiology. Images were in DICOM format and reviewed and measured with the RadAnt™ DICOM viewer (Medixant, Poznań, Poland) software. Cerebral vessel diameters were measured bilaterally at specific locations on maximally magnified images, with consistent measurement locations maintained across serial imaging studies (**Fig. 2B)**. Vessel diameter on each serial image was normalized to the initial angiographic test, with comparisons made only between matching modalities and accounting for anatomical variations or imaging limitations. Vasospasm was defined for each vessel compared with baseline as none/mild if <25%, moderate if 25-50%, and severe if >50%, as previously described^18^. Vessel caliber was also analyzed over time by treatment group. See **Supplemental Materials** for a detailed description of image analysis techniques.

**Figure 2.**
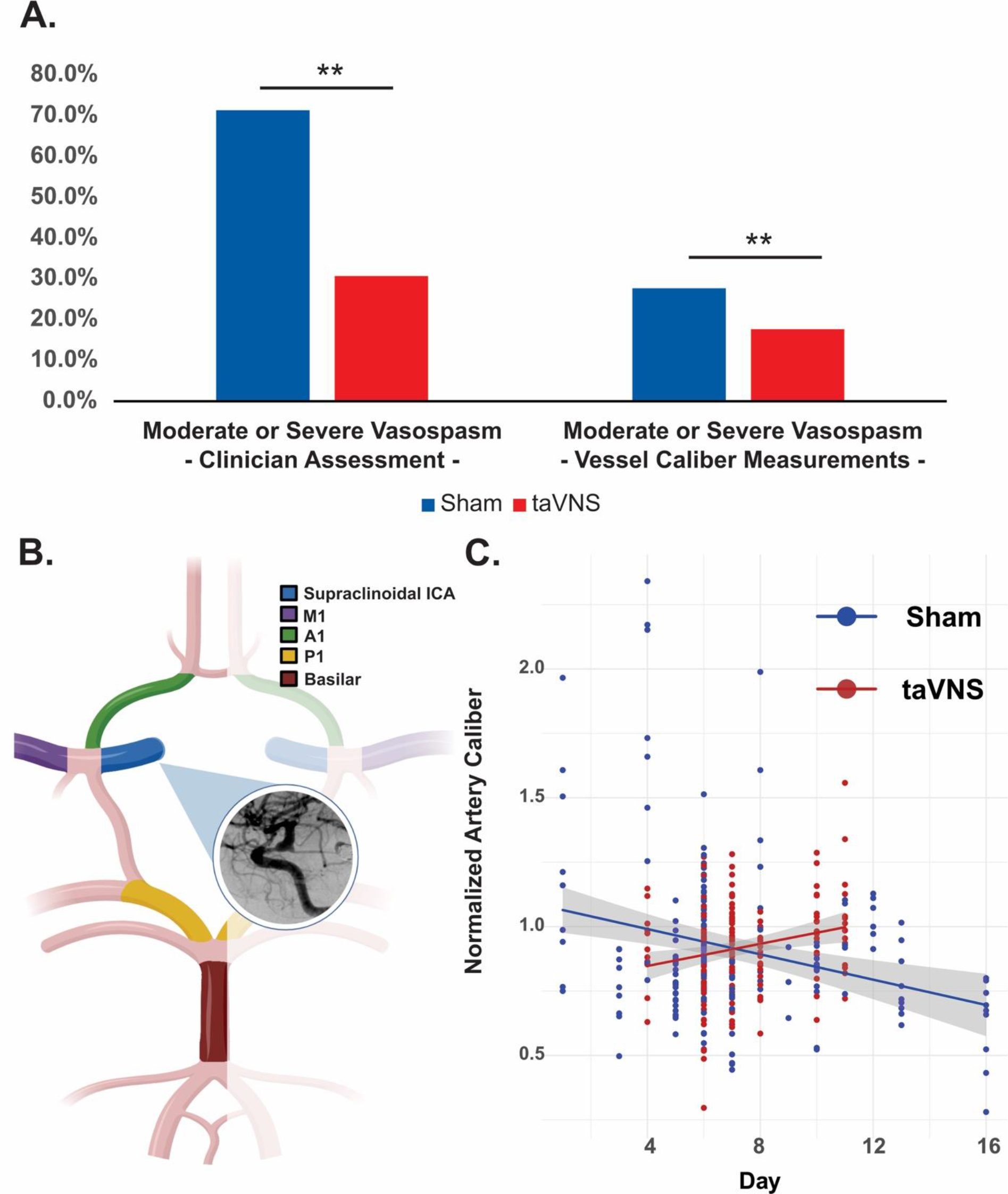
Impact of vagus nerve stimulation on radiographic vasospasm. Rate of radiographic vasospasm, as assessed by a blinded interpreter (left) and serial vessel caliber measurements (right) (**A**). Vessel measurements from a predetermined list of vessel locations (**B**), were used to assess vessel caliber on serial vascular imaging studies (**C**). **p<0.05 ICA= internal carotid artery, taVNS= transauricular vagus nerve stimulation

Secondary outcomes also included the number of vascular imaging studies obtained and interventions undertaken to treat vasospasm, including 1) blood pressure augmentation while in the intensive care unit, 2) treatments performed during catheter angiogram, such as administration of intraarterial vasodilators, 3) the use of intrathecal vasodilators. Presence of infarct on subsequent CT or MRI was also recorded, as a surrogate marker for delayed cerebral ischemia.

### Secondary Outcomes

Additional secondary outcomes included clinical outcome metrics, including discharge destination (home, inpatient rehabilitation facility, skilled nursing facility, hospice, or death), and mRS scores at discharge and first follow-up.

### Sample Size

The estimated required sample size for the pilot NAVSaH clinical trial, as previously published^14^ was 50 patients. However, enrollment to study this endpoint was terminated early given the effect size for the primary aim assessed (presence of moderate/severe radiographic vasospasm) was larger than predicted based on limited preliminary data. Therefore, results are reported based on 27 patients who completed their initial hospital stay and at least their first outpatient follow-up.

### Statistical Methods

Patient demographics and clinical characteristics were summarized using counts and frequencies for categorical variables or means and standard deviations for continuous variables. The distributions of baseline patient characteristics across taVNS and Sham stimulation groups were compared using Student t-test or Chi-square test as appropriate. Between-group differences in the cross-sectional outcomes (such as the presence of moderate or severe radiographic vasospasm, discharge destination, etc.) were compared using a t-test or Chi-square test as appropriate. Between-group differences in these repeatedly measured outcomes (such as modified Rankin Scale (mRS, coded as mRS<3 or not), biomarkers of cytokines, etc.) were compared using linear mixed models (for normality data) or generalized linear mixed models (for non-normality data) to account for potential correlation among multiple measures taken from the same patients. The fixed effects of the model included intervention groups (taVNS vs. Sham), the measurement times (treated as a categorical variable), and their interaction. The random effect included subject-specific intercept which described the average deviation of an individual from the overall level at baseline. One-sided post-hoc tests were performed to assess whether taVNS improved outcomes at specific time points. Normalized vessel caliber was also assessed using a similar mixed model to estimate its average change over time and to compare effect of taVNS on vasospasm, where the measurement time was treated as a continuous variable instead. The assumption of normality was assessed graphically based on residuals out of models. All the analyses were performed using SAS 9.4 (SAS Institutes, Cary, NC). Unless stated otherwise, all the statistical tests were one-sided for outcome variables and two-tailed for baseline characteristics, with a p-value of < 0.05 for significance.

## Results

### Subject Enrollment, Allocation, and Baseline Demographics

#### Participant Flow

A flow diagram detailing enrollment, allocation, and follow up can be found in **Supplemental Fig 1**. Sixty-four patients were assessed for eligibility for enrollment in the trial between 01/2021 and 10/2023. Of those, 27 met inclusion criteria and were randomized to a treatment arm (13 taVNS and 14 patients sham). One patient in each treatment arm ceased treatment sessions and laboratory draws before the completion of the intervention. Overall, 27 patients were analyzed for clinical outcomes based on intention-to-treat, and 26 patients were analyzed for laboratory inflammatory cytokine endpoints, with one patient in the taVNS group excluded from analysis due to only a single baseline lab draw being performed.

#### Baseline data

Patients in both treatment arms were not significantly different with regard to gender, race, Hunt & Hess Score, modified Fisher Scale Score (**Fig. 1 E&F**), or Glasgow Coma Scale (GCS) on admission (**Supplemental Table 1**).

### Safety of Intervention

There were no reportable adverse events, including no reports of pain or irritation related to the stimulation site.

### Impact of taVNS on Radiographic Vasospasm

When interpreted by blinded evaluators, patients treated with taVNS had a significant reduction in the presence of any radiographic vasospasm (p=0.035), or radiographic vasospasm described as moderate or severe (p=0.018) (**Table 1**, **Fig. 2A)**. When cerebral vessel diameter on serial vascular studies were measured, there was also a significant reduction in the number of vessels with moderate or severe vasospasm when normalized to baseline (*p=0.015*) in the taVNS treated goup (**Table 1**, **Fig.2A)**. The normalized vessel calibers significantly increased over time in the taVNS group (with average daily change= 0.030, 95% CI = 0.012 - 0.047), while the average daily change in the sham group is -0.002 (95% CI = [-0.013, 0.008]) (**Fig. 2C**). The serial vascular studies with normalized vessel caliber showed a significant interaction effect between day and treatment (p = 0.0026) (**Fig. 2C).** The mean number of follow up vascular studies obtained was 2.8 (standard deviation = 0.8) for the taVNS group and 3.4 (standard deviation = 1.2) for the sham group. There were similar rates of interventions performed for vasospasm, including blood pressure augmentation, intraarterial vasodilator infusion or angioplasty during angiography, and intrathecal vasodilator administration (**Table 1**).

**Table 1:** Vasospasm following Subarachnoid Hemorrhage.

|  | taVNS | Sham | P-value |
| --- | --- | --- | --- |
| Clinician Assessment of Radiographic Vasospasm* | Number of patients (%) |  |  |
| Any radiographic vasospasm | 7 (53.9%) | 12 (85.7%) | **0.035** |
| Moderate or severe vasospasm on any vascular imaging study | 4 (30.8%) | 10 (71.4%) | **0.018** |
| Quantitative Vasospasm Assessment* | Number of vessels (%) |  |  |
| None/Mild (<25%) | 135 (82.3%) | 136 (72.3%) | **0.028** |
| Moderate (25-50%) | 27 (16.5%) | 45 (23.9%) |  |
| Severe (>50%) | 2 (1.2%) | 7 (3.7%) |  |
| Moderate or Severe Vasospasm | 29 (17.7%) | 52 (27.7%) | **0.015** |
| Vasospasm Intervention† | Number of patients (%) |  |  |
| Blood pressure augmentation | 3 (23.1%) | 6 (42.9%) | 0.41 |
| Intraarterial infusion during angiography | 4 (30.8%) | 6 (42.9%) | 0.35 |
| Angioplasty during angiography | 0 (0%) | 1 (7.1%) | 0.50 |
| Intrathecal vasodilator | 2 (15.4%) | 2 (14.3%) | 0.47 |
|  | Mean (SD) |  |  |
| Number of imaging studies | 2.8 (0.8) | 3.4 (1.2) | 0.28 |
| Number of days of blood pressure augmentation | 1.8 (3.5) | 3.6 (5.1) | 0.28 |
| Delayed Cerebral Ischemia† | Number of patients (%) |  |  |
| Presence of infarct on CT or MRI | 3 (23.1%) | 6 (42.9%) | 0.41 |

P-value is calculated by ANOVA for numerical covariates and chi-square test for categorical covariates
\* one-tailed statistical tests
† two-tailed statistical tests

### Impact of taVNS on Inflammatory Markers following SAH

A total of 26 patients were included in the analysis of plasma TNF-α and IL-6, with one patient excluded due to only having a single baseline level collected. The pro-inflammatory cytokine TNF-α was significantly reduced in the plasma on treatment days 7 and 10 in patients treated with taVNS (p=0.015 and p=0.030, respectively), while IL-6 was significantly lower in the plasma on day 4 in patients treated with taVNS (p=0.024) (**Fig. 3 A&B)**. A total of 14 patients had an extraventricular drain in place and were included in the analysis of CSF TNF-α and IL-6. The pro-inflammatory cytokines TNF-α and IL-6 were both significantly reduced in the CSF on treatment day 13 in patients treated with taVNS (p=0.031 and p=0.025, respectively) (**Fig. C&D)**.

**Figure 3.**
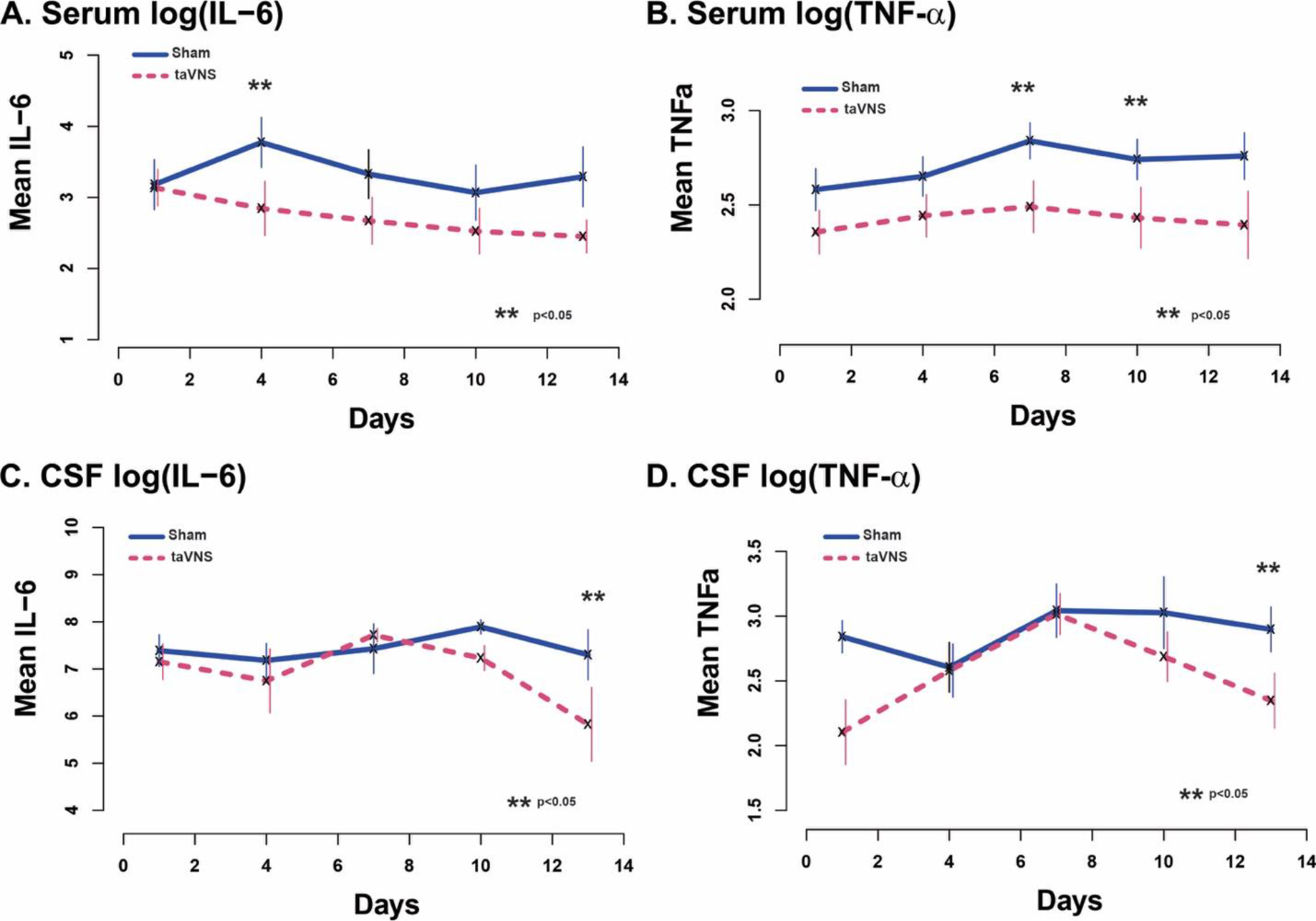
Impact of vagus nerve stimulation on inflammatory cytokines. Serum (**A, B**) and cerebrospinal fluid (**C, D**) measurements of IL-6 and TNF-α over the course of admission following subarachnoid hemorrhage in pg/mL. Day 1 represents baseline prior initiation of first treatment. **p<0.05 taVNS= transauricular vagus nerve stimulation

### Impact of taVNS on Patient Outcomes

Blinded assessments of the mRS scores on admission, discharge, and at first outpatient follow-up are detailed in **Table 2** and **Fig. 4**. One patient in each of the taVNS and sham groups was lost to follow-up, with no mRS assessed after discharge. mRS scores on admission were not significantly different between groups. A good outcome, as assessed via mRS, refers to scores 0- 2, while a poor outcome refers to scores 3-6. The change in mRS over the course of admission and at first follow-up for all patients is illustrated in **Fig. 4C**. Patients receiving taVNS demonstrated higher rates of favorable outcomes compared to those in the sham group both at discharge (38.4% vs 21.4%) and at first follow-up (76.9% vs 57.1%), with a significant difference in improvement observed in the taVNS group from admission to first follow-up (p=0.014), while no significant difference was noted in the sham group (p=0.18)(**Fig. 4D)**. Following hospitalization for SAH, patients had one of 5 discharge outcomes: home, inpatient rehabilitation facility, skilled nursing facility, hospice, or death. No patient in either treatment arm died before discharge from the hospital. With discharge destinations divided into “good” discharge (home and inpatient rehabilitation) versus “poor” discharge (skilled nursing facility, hospice, or death), there was a significantly lower rate of poor discharge in patients treated with taVNS (p=0.04), (**Table 3** and **Fig. 3E)**.

**Figure 4.**
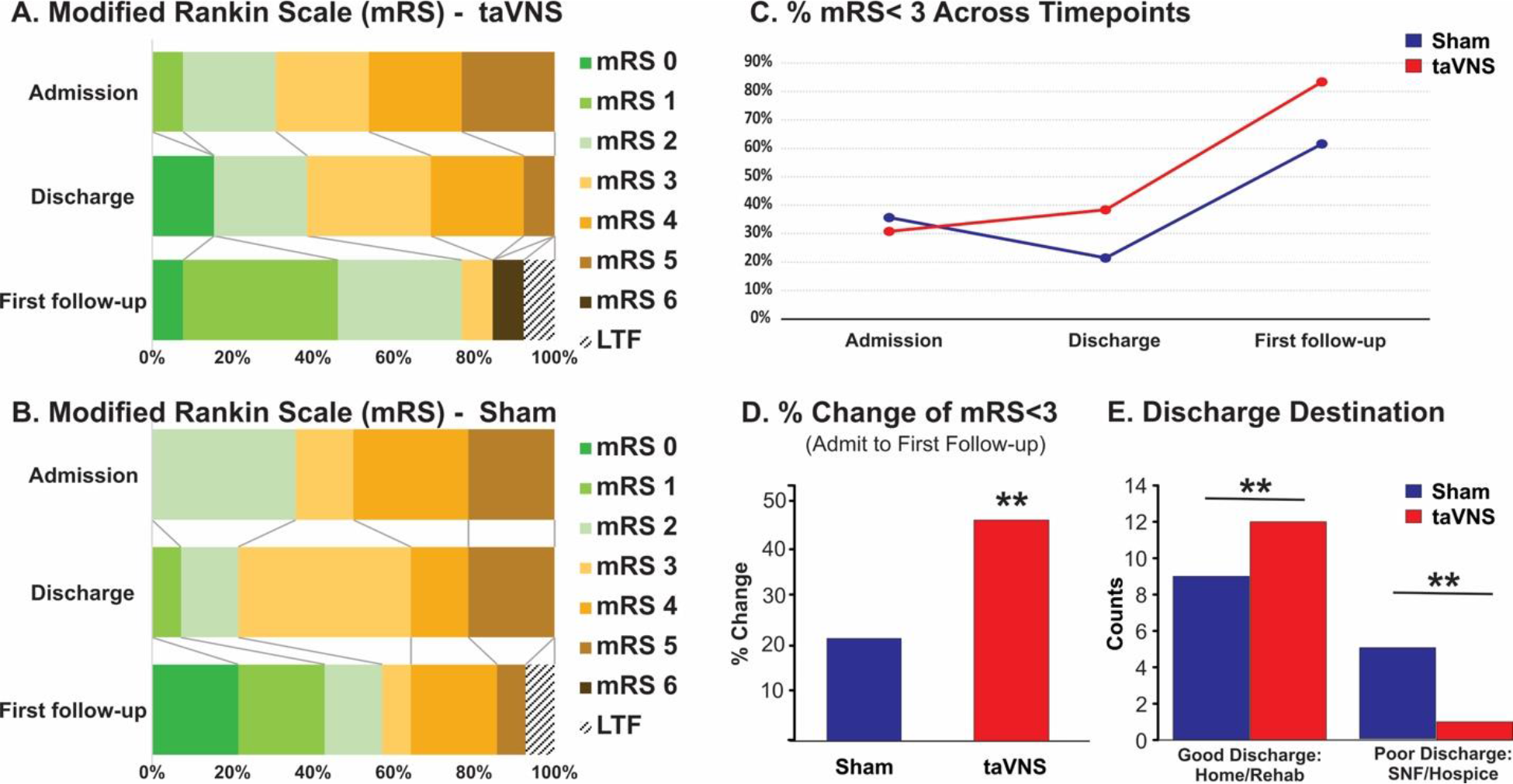
Impact of vagus nerve stimulation on clinical outcomes. Modified Rankin Scale (mRS) scores from blinded assessors at admission, discharge, and at first follow-up for patients treated with VNS (**A**), and sham (**B**) stimulation. Patients with a “good” mRS of <3 shown over time (**C**) and as a change for presentation (**D**) for both treatment groups. Discharge destination by treatment group (**E**) **p<0.05 LTF= lost to follow-up, mRS= modified Rankin Scale, taVNS= transauricular vagus nerve stimulation

**Table 2:** Clinical Outcomes Following Subarachnoid Hemorrhage.

|  | taVNS | Sham | P-value |
| --- | --- | --- | --- |
|  | Number (%) |  |  |
| Discharge destination |  |  |  |
| *Home* | 8 (61.5%) | 6 (42.9%) |  |
| *Inpatient rehabilitation* | 4 (30.8%) | 3 (21.4%) |  |
| *Skilled nursing facility* | 0 (0%) | 5 (35.7%) |  |
| *Hospice* | 1 (7.7%) | 0 (0%) |  |
| *Death* | 0 (0%) | 0 (0%) |  |
| “Good” vs “Poor” discharge destination* |  |  |  |
| “*Good*” discharge | 12 (92.3%) | 9 (64.3%) |  |
| “*Poor*” discharge | 1 (7.7%) | 5 (35.7%) | **0.04** |
| Modified Rankin Scale (mRS) Score | Mean (SD) |  |  |
| mRS on discharge | 2.7 (1.5) | 3.3 (1.2) | 0.18 |
| mRS at first follow-up | 1.8 (1.5) | 2.1 (1.8) | 0.40 |
| Good vs Poor outcome per mRS<sup>†</sup> | Number (%) |  |  |
| Good outcome mRS at discharge | 5 (38.4%) | 3 (21.4%) | 0.21 |
| Good outcome mRS at first follow-up | 10 (76.9%) | 8 (57.1%) | 0.19 |

\* Good discharge destination includes home or inpatient rehabilitation, while poor discharge destination includes skilled nursing facility, hospice, or death
<sup>†</sup> Good outcome is mRS <3
P-value is calculated by ANOVA for numerical covariates and chi-square test for categorical covariates, all one-tailed tests

## Discussion

This study is the first to use non-invasive neuromodulation to mitigate inflammation in SAH patients. Despite improvements in managing SAH patients, mortality and morbidity remain high^1^. Targeting post-hemorrhage inflammation is important for improving outcomes^3,4,6^. We show that taVNS can significantly reduce key inflammatory cytokines (TNF-α and IL-6), reduce cerebral vasospasm, and improve clinical outcomes. These findings provide a new therapeutic approach for treating SAH sequelae.

Following SAH, blood within the subarachnoid space triggers central and systemic inflammatory responses. Key drivers of SAH-induced inflammation are the cytokines TNF-α and IL-6. In animal models and humans, these cytokines are associated with vasospasm, hydrocephalus, delayed cerebral ischemia, and poor outcomes^4,5,19–24^. Numerous anti-inflammatory interventions have been trialed in humans following SAH to target inflammatory pathways. In smaller enrollment studies, there has been some early evidence of clinical benefit with Cyclosporine A^25^ and steroids^26,27^. Other medications, such as Clazosentan^28^, Cilostazol^29^, and IL-1 antagonists^30^ demonstrated no impact on overall outcomes. In larger meta-analysis studies, Simvastatin^31^, Aspirin, non-steroidal anti-inflammatory medications, and thienopyrindines^32^ all demonstrated no improvement. Thus, while some pathway-targeted pharmacological approaches have led to changes in secondary outcomes of vasospasm and delayed cerebral ischemia in clinical trials^29,33^, these approaches have ultimately failed to produce an effective intervention that reliably improves functional or neurological outcomes in SAH patients. These findings, in aggregate, suggest that a narrow, molecular pathway-oriented approach may be insufficient to address the broad cascade of physiologic drivers that result in poor clinical outcomes following SAH.

VNS may provide a broader immunomodulatory approach. VNS reduces inflammation through the “cholinergic anti-inflammatory pathway.” Sensory neurons activated by infection or injury travel to the brainstem via the vagus nerve^34^. Subsequent neural efferent input to the spleen and other organs leads to the release of acetylcholine, which interacts with α7 nicotinic acetylcholine receptors on immunocompetent cells, inhibiting cytokine release in macrophages^35^. VNS has been successfully used in models of cerebral ischemia/reperfusion^36^, rheumatoid arthritis^10^, sepsis^11^, inflammatory bowel diseases^12^, and cerebral aneurysms and SAH^37^. Clinically, VNS has historically been performed by surgical cervical neck dissection and placement of a cuff electrode directly around the nerve. Alternatively, VNS can be accomplished non-invasively by stimulating the auricular branch of the vagus nerve as it courses through the external ear^13^ (**Figure 1**). This non-invasive and low-risk form factor lends itself to deployment in ICU patients who are medically fragile and would not tolerate surgical implantation of a device. Prior to this study, however, there has been a dearth of understanding of VNS effects on SAH patients.

This prospective, randomized trial is the first to report the impact of non-invasive VNS applied following spontaneous SAH. We demonstrate a significant reduction in radiographic vasospasm as assessed by two key metrics: assessment of overall clinical vasospasm by blinded radiology interpreters and quantitative serial vessel caliber measurements. The blinded assessor’s interpretation of vasospasm captures the qualitative radiologic assessment that accounts for large proximal vessel caliber changes, as well as harder-to-quantify distal perfusion alterations from medium vessel caliber changes. The serial quantitative analysis more analytically compares changes in the large proximal vessels over time, limiting potential differences in qualitative assessments by different neuroradiologists. Our finding of significant changes in radiographic vasospasm is a substantial finding, but alteration in radiographic vasospasm alone has not historically been directly or clearly related to changes in functional outcome^38^. Thus, the significant reduction in “poor” discharge destinations (skilled nursing facilities and hospice) and early evidence of superior improvement in mRS between admission and first follow-up implies that the impact of taVNS is not limited to improvement in radiographic vasospasm alone.

In this study, we hypothesized that the underlying mechanism for this vascular and clinical effect was due to taVNS’s ability to blunt the deleterious inflammatory response following SAH. We demonstrate that taVNS can achieve significant reductions in TNF-α and IL-6 both centrally and systemically. Notably, these effects seem to have different time scales. While serum markers generally showed a more linear trend of reduced TNF-α and IL-6 from the onset of treatment, the difference in CSF TNF-α and IL-6 was more delayed and significantly different at day 14. These findings suggest that the serial interventions (i.e., 20 min, twice daily, for 14 days) may have a cumulative effect over time that is necessary to achieve the full clinical benefit. While inflammation is being altered with taVNS, there may be other mechanisms that play a role. Following SAH, there is a widespread sympathetic response, which can manifest with increased risk for cerebral vasospasm, delayed cortical ischemia, and extracerebral organ damage^39^. VNS may restore the SAH-induced autonomic imbalance by enhancing parasympathetic input^40,41^. Another phenomenon observed in SAH patients is cortical spreading depolarizations (CSDs), with evidence of a causal relationship between CSDs and worse neurological outcomes^42^. VNS has been demonstrated to decrease CSDs in a study of migraine^43^, which supports the possibility that taVNS following SAH may have a similar effect. These multi-pathway mechanisms of taVNS may explain the robust clinical benefits seen in this study, while narrowly focused pharmacologic approaches have thus far been unsuccessful.

The results of this study are encouraging, but the current report has limitations. While this study was randomized, it was done at a single institution, and thus, we cannot infer yet whether these findings are more generalizable. This randomized cohort, while having a positive and robust effect, is still small. Ultimately, a larger multi-center clinical trial will be essential to fully validate the clinical effect of taVNS and demonstrate that the treatment can be widely and effectively applied.

## Conclusions

This single-center randomized clinical trial represents the first reported effect of taVNS for the treatment of SAH. Transauricular vagus nerve stimulation modulates the endogenous neuro- immunologic system through a non-pharmacologic bioelectric approach and attenuates the inflammatory response and concomitant vasculopathic sequelae following SAH. Given the low risk, ease of application, and evidence of clinical benefit, this is a novel technique that can potentially impact SAH management in the future.

## Data Availability

All data is available upon reasonable request

## Acknowledgements

Bursky Center for Human Immunology & Immunotherapy Programs (CHiiPs) Core

